# Thyroid dysfunction diagnosis from routine laboratory tests based on machine learning

**DOI:** 10.1101/2021.03.30.21254605

**Authors:** Min Hu, Chikashi Asami, Hiroshi Iwakura, Yasuyo Nakajima, Ryousuke Sema, Tsuyoshi Kikuchi, Tsuyoshi Miyata, Koji Sakamaki, Takumi Kudo, Masanobu Yamada, Takashi Akamizu, Yasubumi Sakakibara

**Affiliations:** AI Strategy Team, Cosmic Corporation Co., Ltd., Tokyo, Japan; First Department of Internal Medicine, Wakayama Medical University Hospital, Wakayama, Japan; Department of Internal Medicine, Gunma University Hospital, Gunma, Japan; Health Care Center, Hidaka Hospital, Gunma, Japan; Department of Internal Medicine, Kuma Hospital, Hyogo, Japan; Department of Biosciences and Informatics, Keio University, Kanagawa, Japan

**Author notes:** **Corresponding Authors** Min Hu, Koishikawa 2-7-3 Tomisaka Building, Bunkyo-ku, Tokyo, Japan, Yasubumi Sakakibara, 3-14-1 Hiyoshi, Kohoku-ku, Yokohama, 223-8522, Japan. These authors contributed equally to this work (Min Hu, Chikashi Asami).

## Abstract

Approximately 2.4 million patients in Japan need treatment for thyroid disease, including Graves’ disease and Hashimoto’s disease. However, only 450,000 of them are receiving treatment, and many patients with thyroid dysfunction remain largely overlooked. In this retrospective study, we aimed to screen patients with hyperthyroidism and hypothyroidism who would greatly benefit from prompt medical treatment. We examined routine laboratory finding data and machine learning algorithms to investigate whether such accurate and robust screening is possible to prevent overlooking and misdiagnosing thyroid dysfunction. We succeeded in developing a machine learning method to construct a classification model for detecting hyperthyroidism and hypothyroidism in patients using 11 routine laboratory tests. We collected electronic health records and medical checkup data from four hospitals in Japan. As a result of cross-validation and external evaluation, we achieved a high classification accuracy for the hyperthyroidism and hypothyroidism models.

## Introduction

Thyroid dysfunction is a leading endocrine disorder with major health implications, including an increased risk of heart disease and hypercholesterolemia. One of the greatest challenges in thyroid dysfunction treatment is to prevent overlooking and misdiagnosing these diseases. Thyroid hormone excess and deficiency are frequently misunderstood and are too often overlooked and misdiagnosed^1^. For hyperthyroidism, the diagnosis may be delayed or missed because some symptoms can be easily attributed to other conditions, such as stress^2^, and are often mistaken for cardiac disease or gastrointestinal malignancies. Hypothyroidism can present with nonspecific constitutional and neuropsychiatric complaints, and patients with hypothyroidism are often misdiagnosed with dementia, cardiac disease, liver disease, or hyperlipidemia and hence not given the proper treatment^3^. The American Association of Clinical Endocrinologists has estimated that in the United States, approximately 4.78% of the population has misdiagnosed thyroid dysfunction^4^ the authors argue that approximately 15 million adults are calculated to have unrecognized thyroid disease^5^. In Japan, it is estimated that approximately 2.4 million patients need treatment for thyroid disease^6^. However, only approximately 450,000 of them are receiving treatment. Thus, patients with thyroid dysfunction are frequently overlooked and misdiagnosed^6,7^.

Hyperthyroidism is a condition that occurs due to excessive production of thyroid hormones. The first step to diagnose hyperthyroidism is to measure free thyroxine (FT4) and free triiodothyronine (FT3) thyroid hormones and thyroid-stimulating hormone (TSH)^6^. In contrast, hypothyroidism is a condition in which serum thyroid hormones decrease. Typical diseases involving hypothyroidism include Hashimoto’s disease and are diagnosed by anti-thyroid antibody tests such as anti-thyroid peroxidase antibody (TPO) and anti-thyroglobulin antibody (TgAb)^5^. Despite their clinical significance, thyroid function tests and anti-thyroid antibody tests are not included in the Japanese national health checkups.

As popular and effective approaches to predictive analytics, machine learning is highly regarded due to its success in diagnosis, prediction, and choice of treatment. Recently, an emerging technique in the field of medical informatics has employed machine learning to accurately derive insights from medical records to support clinical screening and to predict misdiagnosed disease^8^. For instance, a study emphasized the superiority of machine learning technology for predicting cardiovascular risk from routine clinical data^9^. In another study, the incidence of myocardial infarction or cerebral infarction was predicted using the results of a health checkup^10^. Numerous studies have also attempted to assess the efficacy of detecting misdiagnosed diseases, including thyroid dysfunction^11-17^. Aoki et al.^16,17^ found that there were strong, multiple correlations between a set of routine clinical parameters and FT4 in patients with both overt hyperthyroidism and overt hypothyroidism. These studies used pattern recognition methods such as neural networks and predicted the likelihood of thyroid dysfunction from a set of routine clinical tests.

Despite such great efforts, there are still several concerns regarding machine learning applications in the diagnosis of disease. These include the issues of data cleansing, missing value completion, dysfunction labeling criteria, the integration of multiple hospital datasets, and the validation and interpretation of machine learning models. In this study, we developed an explainable artificial intelligence diagnosis support system using machine learning algorithms to identify thyroid dysfunction with routine clinical data to improve medical screening and to prevent overlooking and misdiagnosing thyroid dysfunction. Our study addresses those concerns regarding machine learning applications and provides some possible solutions.

We devised two criteria for dysfunction labeling of data: a thyroid test criterion and a prescription criterion. The thyroid test criterion, which includes the thyroid function parameters TSH and FT4, can be used to clearly model overt and subclinical thyroid dysfunction. However, both TSH and FT4 tests are required; therefore, the number of available data tends to be smaller. More data are available through the prescription criterion, which is based on the presence or absence of doctors’ prescriptions, though this can lead to a problem of confounding overt and subclinical thyroid dysfunction with euthyroidism. Second, we integrated data from four hospitals, including electronic medical records from Wakayama Medical University Hospital, Gunma University Hospital, and Kuma Hospital and annual medical checkup data from Hidaka Hospital. Among the four hospitals, a machine learning model was trained and evaluated via cross-validation by combining patient data from Wakayama Medical University Hospital and Gunma University Hospital with medical checkup data of healthy individuals from Hidaka Hospital. Furthermore, electronic medical record data from Kuma Hospital were used as the external evaluation for the trained models. Third, we examined four typical machine learning algorithms for structured data: gradient boosting decision trees, support vector machines and neural networks used in related studies, as well as logistic regression, which is a common tool in medical studies. Fourth, in terms of the input feature used in machine learning models, features including aspartate aminotransferase (AST), alanine aminotransferase (ALT), gamma-glutamyl transpeptidase(γ-GTP), total cholesterol, hemoglobin, red blood cell count (RBC), creatinine, and sex were selected from the health checkup test list specific to Japan. Alkaline phosphatase (ALP), uric acid (UA), and the UA-to-serum creatinine (S-Cr) ratio were further added, and hence, a total of 11 features were used. To further verify the performance of the models depending on the set of input features, we trained and evaluated models limited to five routine tests, including AST, ALT, γ-GTP, total cholesterol, and sex. Finally, all 24 laboratory findings available in this study were also applied and validated.

## Results

### Model Validation

Table 1-A and Table 1-B are the summaries of the performance results of the machine learning model constructed in this study. As the result of 10-fold cross-validation, as shown in No. I of Table 1-A, the best classification model for overt hyperthyroidism achieved an AUROC = 92.4%, sensitivity = 83.3%, and specificity = 90.9%. The best classification model for overt hypothyroidism achieved an AUROC = 90.5%, sensitivity = 84.4%, and specificity = 86.4%. In the external evaluation, as shown in No. X of Table 1-B, the classification model for overt hyperthyroidism achieved an AUROC = 96.3%, and the classification model for overt hypothyroidism achieved an AUROC = 92.9%. As shown in No. XI of Table 1-B, the classification model for subclinical hyperthyroidism with Feature set 1 achieved an AUROC = 73.0%, and the classification model for subclinical hypothyroidism achieved an AUROC = 69.1%. As shown in No. XII of Table 1-B, the classification model for subclinical hyperthyroidism with Feature set 3 achieved an AUROC = 73.8%, and the classification model for subclinical hypothyroidism achieved an AUROC = 75.2%.

**Table 1-A.** Results of the validation of different models.

| No. |  | I | II | III | IV | V | VI | VII |
| --- | --- | --- | --- | --- | --- | --- | --- | --- |
| Training | Data labeling | Thyroid function test criterion | Prescription criterion | Thyroid function test criterion |  |  |  |  |
|  | Institution combination | Inst. comb. 1 | Inst. comb. 1 | Inst. comb. 2 | Inst. comb. 3 | Inst. comb. 1 |  |  |
|  | ML algorithm | GBDT | GBDT |  |  | SVM | Logistics regression | ANN |
|  | Input features | Feature set 1 | Feature set 1 |  |  |  |  |  |
| Validation | Labeling criteria | Thyroid function test criterion |  |  |  |  |  |  |
|  | Overt/ Subclinical | Overt |  |  |  |  |  |  |
|  | Institution combination | Inst. comb. 1 | Inst. comb. 1 |  |  |  |  |  |
| Hyper-thyroidism | AUROC | 92.4±3.2% | 88.2±3.2% | 91.4±3.0% | 90.7±4.1% | 85.7±5.2% | 86.1±4.6% | 86.7±4.0% |
|  | Sensitivity | 83.3±9.0% | 77.5±8.5% | 81.9±9.2% | 88.4±7.8% | 70.8±11.9% | 76.4±11.4% | 73.5±9.3% |
|  | Specificity | 90.9±6.8% | 86.4±6.2% | 89.8±5.0% | 80.5±12.5% | 91.4±4.6% | 86.0±5.8% | 87.8±5.6% |
| Hypo-thyroidism | AUROC | 90.5±3.6% | 82.4±5.3% | 85.9±4.9% | 86.5±3.5% | 77.9±4.4% | 81.4±5.7% | 79.8±5.3% |
|  | Sensitivity | 84.4±9.6% | 77.4±8.9% | 83.6±4.6% | 80.1±10.1% | 66.9±14.2% | 79.5±8.8% | 73.5±8.8% |
|  | Specificity | 86.4±5.2% | 77.0±11.7% | 78.3±8.8% | 81.0±9.2% | 78.4±14.9% | 73.1±10.7% | 77.1±8.4% |

The mean and standard deviation for the 10-fold cross-validation are shown in each score.

**Table 1-B.** Results of the validation of different models.

| No. |  | VIII | IX | X | XI |
| --- | --- | --- | --- | --- | --- |
| Training | Data labeling | Thyroid function test criterion |  |  |  |
|  | Institution combination | Inst. comb. 1 |  |  |  |
|  | ML algorithm | GBDT |  |  |  |
|  | Input features | Feature set 2 | Feature set 1 |  | Feature set 3 |
| Validation | Labeling criteria | Thyroid function test criterion |  |  |  |
|  | Overt/<br>Subclinical | Overt |  | Subclinical |  |
|  | Institution combination | Inst. comb. 1 | External | Inst. comb. 1 |  |
| Hyper-thyroidism | AUROC | 87.4±4.2% | 96.3±0.3% | 73.0±5.7% | 73.8±6.3% |
|  | Sensitivity | 82.3±6.0% | 87.7±1.8% | 71.7±12.4% | 78.7±11.2% |
|  | Specificity | 83.9±7.7% | 93.5±1.1% | 66.6±14.0% | 61.6±16.3% |
| Hypo-thyroidism | AUROC | 85.5±3.1% | 92.9±0.7% | 69.1±4.1% | 75.2±3.3% |
|  | Sensitivity | 76.0±7.2% | 87.1±3.3% | 68.2±15.8% | 77.7±10.4% |
|  | Specificity | 82.9±9.0% | 84.2±3.7% | 64.3±16.4% | 64.3±13.6% |

The mean and standard deviation for the 10-fold cross-validation are shown in each score.

The result of comparing different labeling criteria is shown in Nos. I and II of Table 1-A. When the prescription criterion was applied as the labeling criterion, the accuracy of the hyperthyroidism classification model achieved an AUROC = 88.2%, and that of the hypothyroidism classification model achieved an AUROC = 82.4%. On the other hand, as shown in No. I, when the thyroid function test criterion was used, the accuracy of the hyperthyroidism classification model achieved an AUROC = 92.4%, and that of the hypothyroidism classification model achieved an AUROC = 90.5%. The model trained on the thyroid function test criterion data achieved superior performance, which was statistically significant by the Wilcoxon test at a p-value of 0.05. The result of comparing models built on different institution combinations is shown in No. I, III, and IV of Table 1-A, as the highest performance was obtained when institution combination 1 was used as the training set, and the accuracy of the hyperthyroidism classification model achieved an AUROC = 92.4%, while that of the hypothyroidism classification model achieved an AUROC = 90.5%.

Among the four machine learning algorithms used in this study, including the GBDT, the SVM, logistic regression, and the ANN, the highest performance was obtained when the GBDT method was applied, as shown in Nos. I, V, VI, and VII of Table 1-A. The accuracy of the hyperthyroidism classification model achieved an AUROC = 92.4%, while that of the hypothyroidism classification model achieved an AUROC = 90.5%, which were statistically significant at p-value 0.05 by the Wilcoxon test. After comparing the performance of different feature sets, as shown in I of Table 1-A and VIII of Table 1-B, when Feature set 3 was applied, the accuracy of the hyperthyroidism classification model was reduced, with an AUROC = 87.4%, and the performance of the hypothyroidism classification model was reduced, with an AUROC = 85.5%, which shows significant differences by the Wilcoxon test at a p-value of 0.05.

The result of comparing different feature sets is shown in No. I of Table 1-A, Nos. VIII and IX of Table 1-B. Comparing to Feature set 2, models constructed on Feature set 1 and Feature set 3 achieved significantly higher AUROC. However, no significant difference was confirmed between the results of Feature set 1(No. I) and Feature set 3(No. IX), hence Feature set 1 with a smaller number of explainable variables was considered sufficient and used for all other validations in this study.

The model of No. I in Table 3 was evaluated using the external dataset from Kuma Hospital, as shown in No. X of Table 1-B. High classification performance was achieved using the external data: AUROC = 96.3%, sensitivity = 87.7%, and specificity = 93.5% for the hyperthyroidism classification model and AUROC = 92.9%, sensitivity = 75.7%, and specificity = 87.1% for the hypothyroidism classification model. No. XI and XII of Table 1-B show that using Feature set 3 slightly improved the classification performance: for subclinical thyroid dysfunction, AUROC = 73.8%, sensitivity = 78.7%, and specificity =61.6%; for hypothyroidism, AUROC = 75.2%, sensitivity = 59.9%, and specificity = 77.7%. In particular, the significance of the hypothyroidism classification models was statistically confirmed by the Wilcoxon test at a p-value of 0.05.

**Table 2.** Evaluation result obtained without considering crosstalk.

| No. |  | A-1 | A-2 |
| --- | --- | --- | --- |
| Training | Thyroid function test criterion | Overt + subclinical |  |
|  | Negative label setting | Crosstalk non-account |  |
| Validation | Thyroid function test criterion | Overt |  |
|  | Negative label setting | Crosstalk non-account | Crosstalk account |
| Hyperthyroidism | AUROC | 94.9±2.4% | 78.5±3.1% |
| Hypothyroidism | AUROC | 91.3±4.0% | 68.1±3.1% |

The mean and standard deviation for the 10-fold cross-validation are shown for the AUROC scores.

**Table 3.** Summary of the data from each institution.

| Institution | Wakayama Medical University | Gunma University | Hidaka Hospital | Kuma Hospital |
| --- | --- | --- | --- | --- |
| Number of prescriptions | 8,249,286 | 34,561,268 | 23,450 | 61,590 |
| Number of patients | 14,249 | 27,133 | 10,482 | 124,863 |
| Average age | 60.9 | 51.7 | 47.7 | 50.3 |
| Male/female ratio | 1.03 (5,888/5,723) | 0.53 (8,143/15,296) | 1.82 (15,125/8,325) | 0.21 |
| Data period | 2010-2018 | 2004-2019 | 2004-2007 | 2007-2020 |

### Feature Importance

The feature importance of each model was examined using Feature set 1. The left picture of Fig. 1 shows the feature importance of the overt hyperthyroidism classification model and the overt hypothyroidism classification model. The three most important features in the overt hyperthyroidism model were ALP, the UA/S-Cr ratio, and total cholesterol. The three most important features in the overt hypothyroidism model were AST, total cholesterol, and RBC. On the other hand, the right picture of Fig. 1 shows the feature importance of the subclinical hyperthyroidism classification model and the subclinical hypothyroidism classification model. The three most important features in the subclinical hyperthyroidism model were ALP, the UA/S-Cr ratio, and S-Cr, and the three most important features in the subclinical hypothyroidism model were total cholesterol, AST, and RBC. For both overt and subclinical disease, ALP and S-Cr were the top related features in the hyperthyroidism classification model, and total cholesterol, AST, and RBC were the top features in the hypothyroidism classification model.

**Fig. 1.**
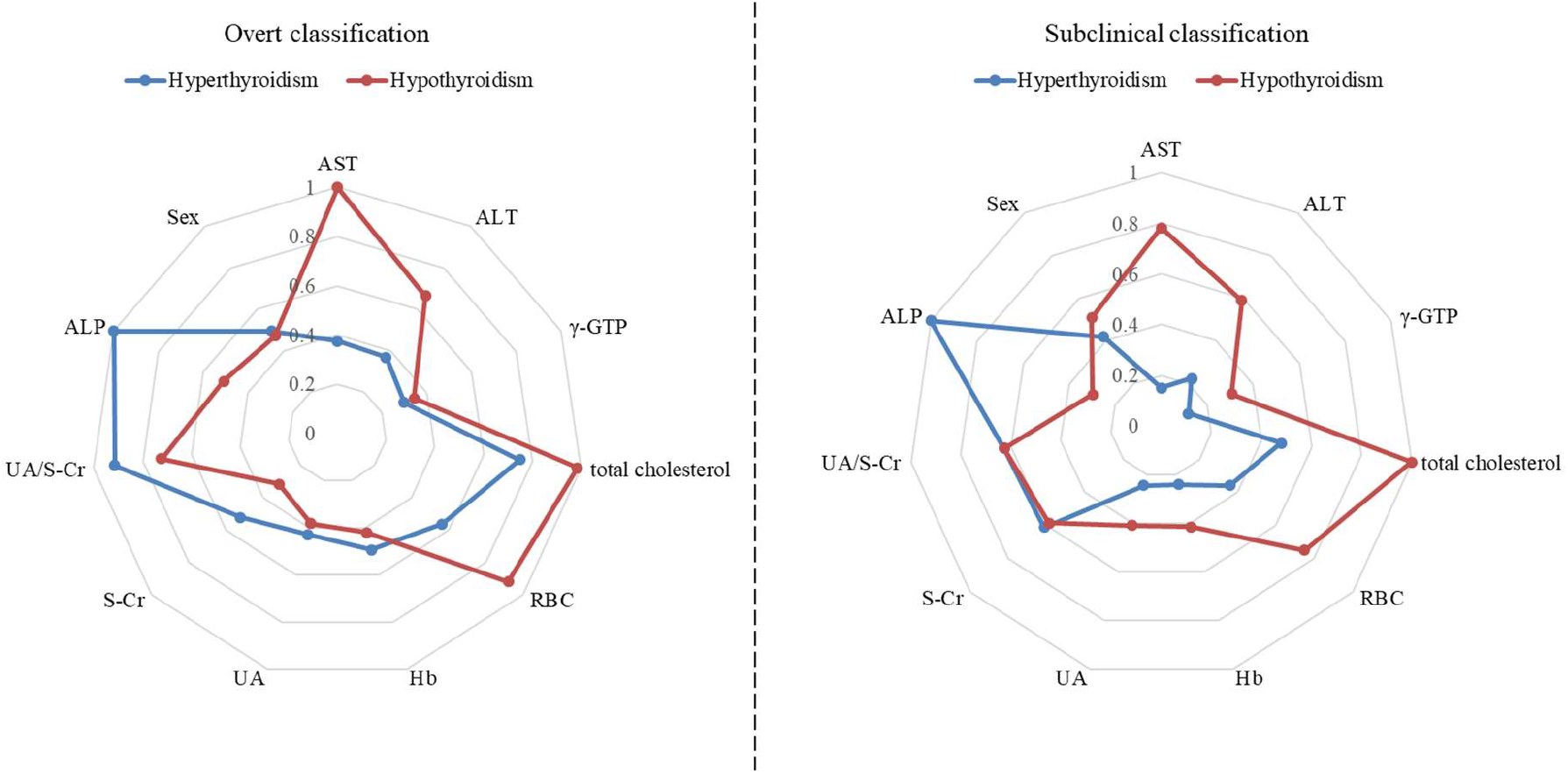
Comparison of feature importance between overt and subclinical thyroid dysfunction classification models.

Furthermore, the importance of features in the subclinical hyperthyroidism and subclinical hypothyroidism classification models using Feature set 3 was determined. As shown in Fig. 2, ALP and the UA/S-Cr ratio were among the three most important features in the subclinical hyperthyroidism classification model when Feature set 1 was used, as well as when Feature set 3 was used. If the five most important features were considered, MCV and MCH, two features added to Feature set 3, were included. These findings suggest that these two features are also likely to be effective in hyperthyroidism classification. On the other hand, as shown on the right side of Fig. 2, a difference was seen in the subclinical hypothyroidism classification model when feature set 1 was used vs. when Feature set 3 was used. The three most important features in the model that used Feature set 1 were total cholesterol, AST, and RBC, whereas the three most important features in the model that used Feature set 3 were total protein, total cholesterol, AST, and the UA/S-Cr ratio. These findings suggest that total protein is likely to be effective in classifying subclinical hypothyroidism.

**Fig. 2.**
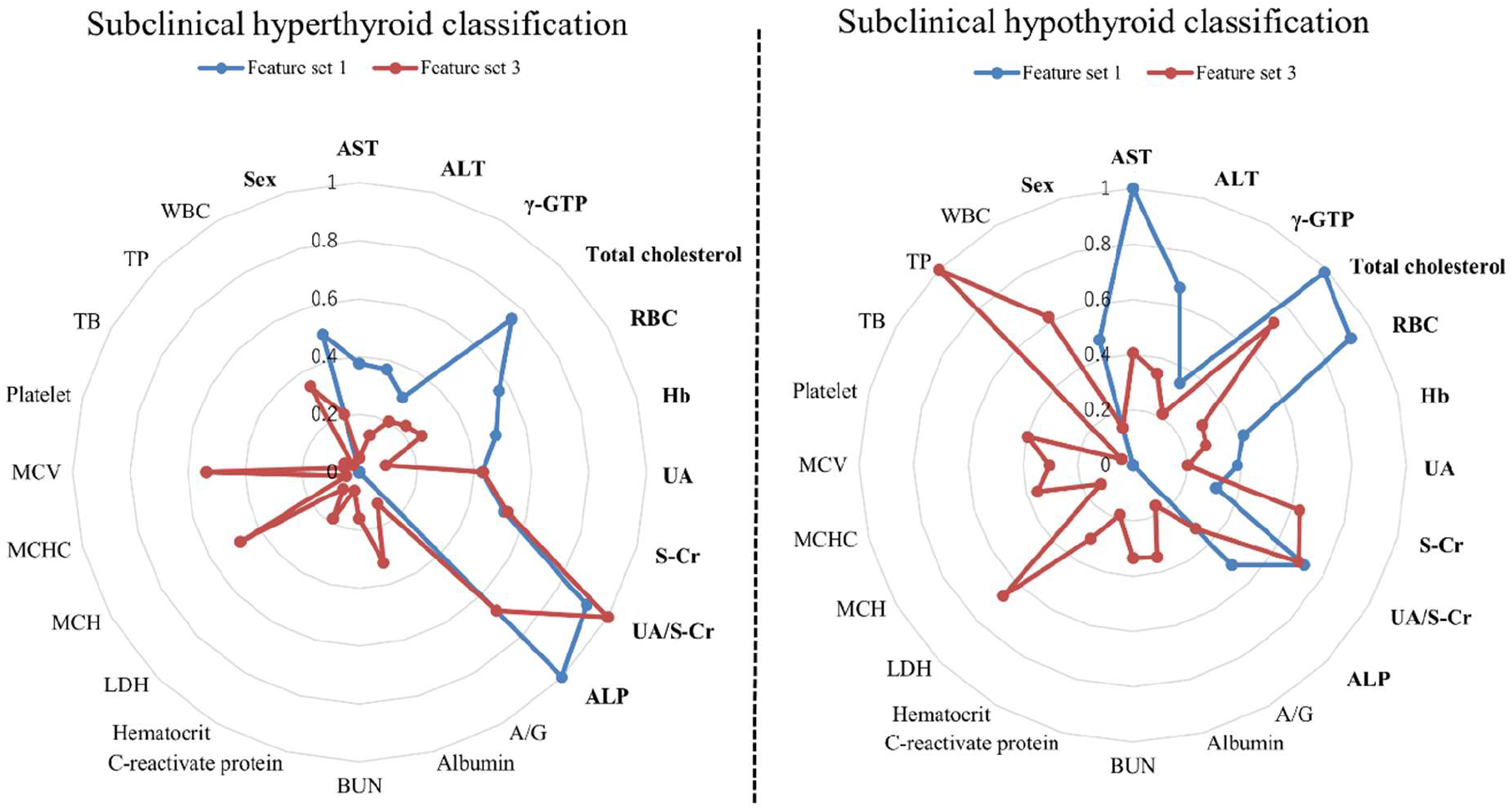
Comparison of feature importance between models built on Feature set 1 and Feature set 3.

## Discussion

### Feature Importance

The correlation of routine laboratory test parameters such as ALP, S-Cr, UA, and RBC with thyroid dysfunction has been pointed out in many previous studies. According to studies on the relationship between thyroid dysfunction and liver function^18,19^, a correlation was confirmed between the increase in ALP and hyperthyroidism, as the ALP value was significantly higher when bone metabolism increased in Graves’ disease, which is a typical disorder of hyperthyroidism. Sönmez^20^ examined data from 433 patients and reported that S-Cr in the hyperthyroidism group was significantly lower than that in the euthyroid group. TSH and S-Cr were also reported to have a significantly negative correlation with overt hypothyroidism^21^. Dorgalaleh et al.^22^ suggested that thyroid dysfunction directly affects most blood values, including RBC, and health professionals must pay attention to such effects. The correlation between hypothyroidism and hyperuricemia has also been confirmed by multiple studies^23,24^.

### Comparison with Related Studies

Several previous studies revealed promising results from the use of machine learning approaches for predicting thyroid dysfunction^16,17^.

Similar to the present study, Aoki’s et al.^17^ study used pattern recognition methods such as neural networks to predict the likelihood of thyroid dysfunction from a set of routine test parameters such as ALP, S-Cr, and TC. Their results suggested that most patients with overt thyroid dysfunction could be screened by using a set of routine clinical data without measuring thyroid hormone levels. The correct rate of 91.3% was reported in the hyperthyroidism classification model, and the correct rate of 90.0% was reported in the hypothyroidism classification model. Their results suggested that there is a high correlation between a set of routine laboratory tests and thyroid dysfunction. However, the model verification of these studies used the leave-one-out method instead of cross-validation and used the correct rate as the indicator instead of the AUROC. Thus, the model evaluation was considered insufficient.

Unlike the present study, one drawback of these previous studies is that they have not considered crosstalk in the data labeling process. For hyperthyroidism classification in this study, the hyperthyroidism group was used as a positive label, and both the control and hypothyroidism groups were negatively labeled. For the hypothyroidism classification in this study, the hypothyroidism group was used as a positive label, whereas both the control and hyperthyroidism groups were negatively labeled (referred to as “crosstalk on”). On the other hand, related studies^16,17^ performed classification by setting thyroid dysfunction patients (with hyperthyroidism or hypothyroidism) as a positive label and only the control group as a negative label (referred to as “crosstalk off”). Therefore, we evaluated the performance of the models with similar settings as these studies. As shown in column A-1 of Table 2, when only the control group was labeled negative in both the training data and the validation data, a high classification performance, with AUROCs = 94.9% and 91.3%, was achieved in the classification of overt hyperthyroidism and overt hypothyroidism, respectively. However, as shown in column A-2 of Table 2, when both the control group and the hypothyroidism group were labeled negative in the validation data of overt hyperthyroidism and when both the control group and the hyperthyroidism group were labeled negative in the validation data of overt hypothyroidism, the classification performance was reduced, with AUROCs = 78.5% and 68.1%, respectively. The classification performance dropped significantly in the models in which crosstalk was not considered during the negative labeling process.

### Limitations

In the current study, subjects on medication may be included in the data extraction process of this study. Though we extracted only the laboratory test parameters at each subject’s first visit to avoid including the influence of thyroid dysfunction treatment, some subjects might have already been on medication before being referred to the hospitals in our study. These subjects on medications may have an unexpected impact on the models we built in this study.

Another limitation of this study is that the hypothyroidism classification models exhibited lower performance than the hyperthyroidism classification models. This result is attributed to differences in the respective serum hormones and underlying molecular mechanisms^25^. The various nonspecific symptoms of hypothyroidism may not manifest simultaneously, resulting in a subclinical rate larger than that of hyperthyroidism. In addition, patients with hypothyroidism, such as those with Hashimoto’s thyroiditis, are dependent upon long-term levothyroxine treatment, which may affect the manifestations of routine laboratory findings.

Furthermore, in the external evaluation of this study, the subclinical classification model showed lower overall results than the overt classification models. Among cases of subclinical thyroid dysfunction, the cause of subclinical hypothyroidism is associated with chronic thyroiditis (Hashimoto’s disease), of which approximately 60-80% of cases are related to thyroid autoantibodies^26^. On the other hand, the causes of subclinical thyrotoxicosis are classified into extrinsic overdose of thyroid hormone drugs and endogenous hyperthyroidism such as Graves’ disease^27^. Most cases of subclinical thyroid dysfunction, such as subclinical thyrotoxicosis and subclinical hypothyroidism, have no subjective symptoms and are usually considered to be transient^28,29^. The performance of the screening method may have been limited because symptoms of subclinical thyroid dysfunction are usually minor compared to those of overt thyroid dysfunction, and the phenotype of subclinical thyroid dysfunction may not be reflected in the results of routine laboratory examinations.

### Conclusion

This study evaluated a screening method to discriminate hyperthyroidism and hypothyroidism from electronic medical records or routine laboratory finding data from health checkups using a machine learning method with the aim of preventing missed diagnosis of thyroid dysfunction. This is a versatile new screening method that was successfully developed from a machine learning model construction method to discriminate patients with hyperthyroidism and those with hypothyroidism using 11 features. High accuracy was achieved in the discrimination of evident hyperthyroidism or hypothyroidism, and although the discrimination accuracy of subclinical hyperthyroidism or hypothyroidism was not satisfactory, these features can be useful for nonspecialists for thyroid diseases.

The quality of life of patients is expected to improve by applying the model developed in this study. If thyroid dysfunction is screened using our method in healthcare facilities, including hospitals and health checkup facilities, prompt and accurate diagnostic support can be provided from only routine laboratory tests.

## Methods

### Data Source

In the present study, we acquired laboratory finding datasets from different clinical university medical institutions in Japan, including Wakayama Medical University Hospital, Gunma University Hospital, Hidaka Hospital, and Kuma Hospital. The anonymized electronic medical records included age, sex, diagnosis codes for insurance billing, prescribed drugs, and biochemical test results. The institutional ethical review boards of the three institutions at which the study was conducted gave their approval.

A sample of 176,727 subjects in total, aged between 13 and 88 and from different regions in Japan between 2004 and 2019, were included in our study, as illustrated in Table 3. Among the four institutions, Wakayama Medical University Hospital and Gunma University Hospital are hospitals affiliated with a medical college, Hidaka Hospital is a regional medical care support hospital, and Kuma Hospital is a hospital specializing in thyroid diseases. The data of the 176,727 subjects consisted of doctor evaluations, prescriptions, clinical examinations, and laboratory findings. The doctor evaluations addressed medical history, medication use, and differential diagnosis, among other topics. If a subject was prescribed medication, the name and dose of the prescription were recorded. The examinations involved anthropometric measurements and laboratory tests, among others. The institutional ethical review boards of the three institutions at which the study was conducted gave their approval (Approval Number of Wakayama Medical University Hospital: 2301, Hidaka Hospital: 257, Gunma University Hospital: HS2018-245)

The K-nearest neighbor (KNN) algorithm was used to predict and complement the missing values, with k set to 3 in the data filling process. A previous study^11^ reported the KNN algorithm to substantially increase the number of applicable subjects. Compared with missing value deletion, the KNN algorithm is easily applied, performs well for nonparametric datasets and provides a larger sample size. Furthermore, since the age and sex distributions were different among the institutions, as shown in Table 3, we also conducted random undersampling to fix the gaps in these differences. From this dataset, the model was constructed using thyroid patient data from Wakayama Medical University and Gunma University and control group data from Hidaka Hospital and was evaluated using cross-validation. To validate the external data, the model was also evaluated on the dataset from Kuma Hospital.

### Construction of the Machine Learning Model

As shown in Table 4, four verification items were devised in this study to improve the performance of our machine learning model. The criteria of data labeling and the combination of multiple institutions were evaluated first. Then, four different machine learning algorithms and three sets of input features were evaluated to achieve the best performance of our thyroid dysfunction classification models.

**Table 4.** List of verification items.

| No. | Verification item | Option |  |  |  |
| --- | --- | --- | --- | --- | --- |
| 1 | Training data labeling | Thyroid function test criterion | Prescription criterion |  |  |
| 2 | Institution combination (for patient data and control group data) | Institution combination 1 (Inst. comb. 1) | Institution combination 2 (Inst. comb. 2) | Institution combination 3 (Inst. comb. 3) | External |
| 3 | Machine learning algorithm | GBDT | SVM | Logistic regression | ANN |
| 4 | Input features | Feature set 1 | Feature set 2 | Feature set 3 |  |

### Data Labeling Criterion

According to the guidelines of the Japanese Thyroid Association for the diagnosis of hyperthyroidism and hypothyroidism, if thyroid disorder is suspected from the clinical findings, first, a thyroid function test (TSH and FT4 measurement) is conducted, and on the basis of these results, thyroid disorder is classified into three categories—hyperthyroidism, hypothyroidism, or euthyroidism^37^. Therefore, we devised and compared the performance of two data labeling criteria.

We first devised the labeling criterion by using the result of the thyroid function test as a reference (hereinafter referred to as the “thyroid function test criterion”). Specifically, in the dataset from Wakayama Medical University, FT4 and TSH were measured with ECLusys kits. TSH < 0.5 and FT4 > 1.7 were defined as overt hyperthyroidism, TSH < 0.5 and 0.9 ≤ FT4 ≤ 1.7 as subclinical hyperthyroidism, TSH > 5.0 and FT4<0.9 as overt hypothyroidism, and TSH > 5.0 and 0.9 ≤ FT4 ≤ 1.7 as subclinical hypothyroidism (TSH unit: μIU/mL; FT4 unit: ng/dL). In the dataset from Gunma University, in which FT4 and TSH were measured with the Architect kits, TSH < 0.35 and FT4 > 1.48 were defined as overt hyperthyroidism, TSH < 0.35 and 0.7 ≤ FT4 ≤ 1.48 as subclinical hyperthyroidism, TSH > 4.94 and FT4 < 0.7 as overt hypothyroidism, and TSH > 4.94 and 0.7 ≤ FT4 ≤ 1.48 as subclinical hypothyroidism. In this study, overt and subclinical hyperthyroid patients were collectively referred to as the hyperthyroidism group, and overt and subclinical hypothyroid patients were collectively referred to as the hypothyroidism group.

Data for the control group were extracted from the third institution, Hidaka Hospital, and consisted of the test results from regular medical examinations. We extracted comprehensive medical examination data for subjects who did not have any symptoms suggesting thyroid dysfunction or abnormal values in the laboratory tests of TSH and FT4. The normal ranges were set to 0.34-3.88 μIU/mL for TSH and 0.95-1.74 ng/dL for FT4. Random undersampling was conducted for the control group in such a way that the sample size of the control group was equivalent to the sizes of the hyperthyroidism and hypothyroidism groups. The thyroid function test criterion required both TSH and FT4 test results, but a small number of patient records had both of these results. Therefore, as an alternative solution, we devised another criterion of labeling the training data according to the presence of a prescription (hereinafter referred to as the “prescription criterion”) for thyroid disorder. Specifically, the use of the prescription criterion satisfies the following conditions: (a) it includes patient records with standard prescribed medications for thyroid dysfunction (including thiamazole, propylthiouracil, and potassium iodide for the hyperthyroidism group and levothyroxine and thyronamine for the hypothyroidism group) obtained at the patients’ first visits, (b) it includes patients not diagnosed with thyroid nodules, (c) it includes patient records containing laboratory findings obtained within four weeks after the patient’s first prescription, and (d) it excludes records with missing values for more than half of our selected features. Since the age distributions were different among the institutions, as shown in Table 2, we also conducted data sampling to fix the gaps in these differences.

In machine learning, a control group is generally used as a negative label. Since hyperthyroidism and hypothyroidism are conditions of thyroid dysfunction, both often express similar symptoms and effects on some routine laboratory findings (e.g., Hb is decreased in both hyperthyroidism and hypothyroidism patients). Therefore, we considered the confounding of hyperthyroidism and hyperthyroidism as “crosstalk” and refined the labeling criteria in such a way that the negative label was set as both the healthy subjects of the control group and the patients of the opposite type of thyroid dysfunction. For instance, in the data labeling process of the hyperthyroidism classification model, the hyperthyroidism group was set as a positive label, whereas both healthy subjects of the control group and hypothyroidism patients were set as a negative label.

### Integrating Multiple Hospital Datasets

The demographics were different among the three institutions from different districts. To investigate the effect of integrating the datasets from these three hospitals, we explored three combinations of datasets to increase the generalization ability of our models. Specifically, three options on datasets, namely, thyroid dysfunction group data from both Wakayama Medical University and Gunma University and control group data from Hidaka Hospital (referred to as Inst. comb. 1), thyroid dysfunction group data from Wakayama Medical University and control group data from Hidaka Hospital (referred to as Inst. comb. 2), and thyroid dysfunction group data from Gunma University and control group data from Hidaka Hospital (referred to as Inst. comb. 3), were set to train and evaluate the models.

### Machine Learning Algorithms

Four representative machine learning algorithms were applied, and their performance in thyroid dysfunction classification was evaluated:

The gradient boosting decision tree (GBDT), as proposed by Friedman^30^, produces a prediction model in the form of an ensemble of weak prediction models, typically decision trees. The GBDT is based on a machine learning technique that consists of an “ensemble” family of algorithms, creates multiple models (called weak learners), and combines them to increase the prediction accuracy. The main idea of this technique is to build a set of decision trees and use them to classify a new case. Each decision tree is generated using randomly selected variable subsets from all feature variables and a randomly selected subset of data combined by bootstrapping^31^. In this study, we employed the most accurate algorithm, called CATBoost^32^, in the GBDT family.

The artificial neural network (ANN) is a well-established classification technique that is widely used in pattern recognition studies. In general, an ANN consists of 3 layers: an input layer that receives information, a hidden layer that processes information, and an output layer that calculates the results^33^. In the present study, a standard feed-forward ANN was applied due to its relative simplicity and stability.

The support vector machine (SVM) is a supervised machine learning technique that is widely used in pattern recognition and classification problems^34^. In the approach of this method, each data sample is a vector whose dimensions are equal to the number of features to be considered, and the SVM creates a hyperplane that separates samples into two categories. The induced hyperplane is constructed to maximize its distance from the samples of both classes.

This algorithm achieves high classification performance by using special nonlinear functions called kernels to transform the input space into a multidimensional space^34^. In this study, the radial basis function kernel is used.

Logistic regression is a statistical classifier that provides the probability for predicting the labeled class of categorical type by using a number of attributes. Logistic regression is frequently used to examine the risk relationship between disease and exposure, with the ability to test for statistical interaction and control for multivariable confirmation^35^. Logistic regression is a linear model and is used as the baseline model for the performance comparison,

### Explanatory Features (Variables) for Machine Learning

Features from a subject’s record were designed to sufficiently explain factors that were related to thyroid dysfunction. We used 5 explanatory variables as the baseline set of features (referred to as Feature set 2): sex, AST, ALT, γ-GTP, and total cholesterol, of which four features were the required laboratory tests conducted in the Japanese national health screening program called Specific Health Checkups. In addition, hemoglobin (Hb), RBC, creatinine (S-Cr), ALP, and UA, which are the tests measured only at the doctor’s discretion, are reported to be highly relevant to thyroid dysfunction^14,15^, these were added to the above items. We also included the UA/S-Cr ratio in this study considering that a reduction in S-Cr has been reported in hyperthyroidism, while UA has not been confirmed to fluctuate with thyroid dysfunction. To discriminate hyperthyroidism from renal dysfunction, which usually leads to an increase in both S-Cr and UA, we introduced the UA/S-Cr ratio as one of the features to improve the classification performance. Eleven tests (referred to as Feature set 1) in total were used as features to train machine learning models in this study.

### Model Validation

Cross-validation was applied to evaluate the performance of our machine learning method in classifying patients. The evaluation was conducted by extracting 9/10 training data and 1/10 test data by conducting 10-fold cross-validation. This was repeated 10 times to extract the training and test data uniformly, and the average and standard deviation of each evaluation score of each time were calculated. During the model training and test process, we avoided including the same subject in both the training dataset and test dataset. The following measures were used for the performance evaluation criteria: the area under the receiver operating characteristic curve (AUROC); area under the precision-recall curve (AUPRC); sensitivity, defined by TP/(TP+FN); and specificity, defined by TN/(TN+FP), where TP is the number of true positives, TN is the number of true negatives, FP is the number of false positives, and FN is the number of false negatives. Note that the cutoff value for classification as positive or negative is determined by the Youden^36^ index. Finally, the AUROC performance difference between models was verified as statistically significant by the Wilcoxon signed-rank test.

In addition, the data of Kuma Hospital were employed for external validation. The model was constructed using the hyperthyroidism group and the hypothyroidism group of Wakayama Medical University and Gunma University and the control group of Hidaka Hospital as the training data. The model was evaluated using the hyperthyroidism group and hypothyroidism group of Kuma Hospital and the control group of Hidaka Hospital (referred to as External).

### Classification of Subclinical Thyroid Dysfunction

In the guidelines of the Japan Thyroid Association^37^, subclinical hypothyroidism is defined when the FT4 level is within the normal limit but the TSH level is higher than normal. On the other hand, subclinical hyperthyroidism is defined when FT4 is within the normal limit but TSH is lower than normal. Compared to overt thyroid dysfunction, where both TSH and FT4 are out of the standard ranges, it is difficult to classify subclinical thyroid dysfunction. This study evaluated the classification performance of the machine learning model by using subclinical standards in the thyroid function test criterion labeling method. We further extended the feature set in an attempt to improve model performance and selected 24 tests (referred to as Feature set 3), which was all the laboratory tests available in this study ^1^.

### Feature Importance

To further understand how each feature contributes to the classification of patients in our model, we introduced feature importance. Feature importance represents the factor by which the model error is increased compared to the original model error. In the decision-tree–based machine learning algorithms, including the GBDT, impurities and the features at which the node is split are recorded for all the nodes when the decision tree learning is finished, and the decision tree calculates the feature importance using this information^31^.

## Data Availability

The data that support the findings of this study are available from the Japan Thyroid Association, but restrictions apply to the availability of these data, which were used under license for the current study and therefore are not publicly available. Data are, however, available from the authors upon reasonable request and with permission from the Japan Thyroid Association.

## Code Availability

The code developed in this project is confidential, but may be obtained with Data Use Agreements with the Japan Thyroid Association. Researchers interested in access to the code should contact MH and CA at. It can take some months to negotiate data use agreements and gain access to the data and the code for data cleaning and analysis. The authors will assist with any reasonable replication attempts for two years following publication.

## Data Availability

The data that support the findings of this study are available from Japan Thyroid Association but restrictions apply to the availability of these data, which were used under license for the current study, and so are not publicly available. Data are however available from the authors upon reasonable request and with permission of Japan Thyroid Association.

## Acknowledgements

This study would not have been possible without the exceptional support of Dr. Akira Miyauchi, who shared insightful comments on this project, provided the opportunity to validate the models on an external dataset and improved this study in innumerable ways. Dr. Masako Akuzawa and Dr. Yoshitaka Ando facilitated this project to access the dataset of the control group in Hidaka Hospital, which significantly improved the generalization performance of the thyroid dysfunction classification models built in this project.

This research received no specific grant from any funding agency in the public, commercial, or not-for-profit sectors.

## Author Contributions

MH and CA implemented the software, analyzed the data, and cowrote the paper. MY and TA supervised the research and collected the medical data. HI, YN, KS, and TK aided in the feature set design and interpretation of the results of the models and collected the medical data. RS and TM contributed to the design of the research and to the writing of the manuscript. YS designed and supervised the research, analyzed the data, and cowrote the paper. All authors read and approved the final manuscript.

## Footnotes

1 Feature set 3 includes sex, AST, ALT, γ-GTP, total cholesterol, RBC, hemoglobin, uric acid, S- Cr, uric acid/S-Cr ratio, ALP, albumin-globulin ratio, albumin, blood urea nitrogen, C-reactive protein, hematocrit, lactate dehydrogenase, mean corpuscular hemoglobin, mean corpuscular hemoglobin concentration, mean corpuscular volume, platelet count, total bilirubin, total protein, and white blood cell count.

